# Comparison of the COVID-19 infection risks by close contact and aerosol transmission

**DOI:** 10.1101/2020.07.13.20152900

**Authors:** Xiaole Zhang, Jing Wang

**Affiliations:** Institute of Environmental Engineering (IfU), ETH Zürich, Zürich, CH-8093, Switzerland; Laboratory for Advanced Analytical Technologies, Empa, Dübendorf, CH-8600, Switzerland

## Abstract

A comprehensive understanding of the transmission routes of the severe acute respiratory syndrome coronavirus 2 (SARS-CoV-2) is of great importance for the effective control of the spread of Corona Virus Disease 2019 (COVID-19). Human-to-human transmission by close contact where large respiratory droplets play a significant role has been established as the main transmission route. At the same time the transmission by small aerosol is getting increasing attention. There is no distinct boundary between droplets and aerosol in nature so it is natural to investigate the infection risk due to aerosol. Here, we utilized a newly developed dose-response relation, combined with a box model for the exposure estimation, to quantitatively evaluate the infection risk of SARS-CoV-2 through aerosol transmission and compared with the risk due to close contact. The results indicated that the median infection risk via aerosol transmission was about 3.7×10^−5^ (95% confidence interval: 3.5×10^−6^ to 4.4×10^−4^) for one hour of exposure in a room with the size of 10 m (width)×10 m (length)×3 m (height) with one infected individual in it. The risk was more than three orders of magnitude lower than the risk at short distance, about 12.8% within 1 m, based on a meta-analysis. A simple exponential regression model *Risk*=10^−0.90×*D*+0.10^ (*D*<=5 m) could be utilized to characterize the magnitude of infection risk in the considered scenario based on the distance *D* from the infected individual. With prolonged exposure duration and large exposed population, the infection caused by aerosol transmission could be considerable, thus it is necessary to be cautious for the potential aerosol transmission risk in such situations.

---

A comprehensive understanding of the transmission routes of the severe acute respiratory syndrome coronavirus 2 (SARS-CoV-2) is of great importance for the effective control of the spread of Corona Virus Disease 2019 (COVID-19). A recent open letter from 239 scientists ^1^ appealed to the relevant communities and organizations to recognize the potential airborne transmission of SARS-CoV-2 through microscopic respiratory particles (aerosols).

A quantitative assessment of the infection risk through different transmission routes is essential to evaluate their relative importance and to prioritize the control measures. A recent study by Chu et al. ^2^ has estimated the infection risk by close contact, which was about 12.8% within 1 m and about 2.6% at further distance through a systematic review and meta-analysis on the betacoronaviruses causing severe acute respiratory syndrome (SARS), Middle East Respiratory Syndrome (MERS) and COVID-19. However, the studies for aerosol transmission risk are still scarce in part due to the lack of dose-response relation.

Recently, we developed a simple framework ^3^ to deduce the dose-response relation for coronaviruses, by integrating the *a priori* dose-response relation for SARS-CoV ^4^ based on mice experiments, the recent data on infection risks from COVID-19, SARS and MERS meta-analysis ^2^ and the viral shedding ^5^. Here, we utilized the newly developed dose-response relation, combined with a box model for the exposure estimation, to quantitatively evaluate the infection risk of SARS-CoV-2 through aerosol transmission.

An exponential model, *p*=1-exp(-*d*/*k*), was adopted for the dose-response relation, where *p* is the infection risk, *d* is the exposure dose, and *k* is the pathogen dependent parameter. It was estimated that *k* was from 6.2×10^4^ to 7.3×10^5^ virus copies dependent on the contributions of the airborne virus-laden particles to the total infection risk ^3^. The value of *k* was about 1.5×10^5^ if the aerosol transmission contributed half of the infection risk. Monte Carlo simulations were conducted to assess the risk. Here, it was assumed that *k* was a stochastic variable following the triangular distribution with lower limit, upper limit and mode of 6.2×10^4^, 7.3×10^5^ and 1.5×10^5^ copies.

To illustrate the relative importance of aerosol transmission compared to that of close contact, we considered a simple scenario of a room with the size of 10 m (width)×10 m (length)×3 m (height). The typical ventilation rate for offices ^6^, 1 ACH (Air Changes per Hour), was adopted for the scenario. One infected individual was assumed to be at the center of the room. The infection risk of close contact from the meta-analysis ^2^ was induced by various durations from any duration to a minimum of 1 h. Representative exposure duration of 1 h was utilize for the development of the dose-response relation. It is close to the total duration of close contact between a nurse / health worker and a patient per day ^7^. One hour of exposure in the room was also adopted for the assessment of the infection risk here.

A box model ^8^ was utilized to estimate the virus concentration and deposition in the room. The box model assumed that the airborne virus-laden particles were well mixed and the virus concentration was homogeneous in the room. A recent study based on computational fluid dynamics ^9^ indicated the aerosol mainly remained below 2 m for indoor environment, so 2 m was utilized as the mixing height instead of the room height (3 m). Four key processes were considered in the model, i.e. viral shedding, biologic decay, ventilation and deposition. Recently, Leung et al. ^5^ has shown that the viral shedding (*E*_*virus*_) was about 10^2^ to 10^5^ virus copies with a geometric mean of about 10^4^ copies in the samples of respiratory particles from the symptomatic individuals infected by coronavirus (NL63, OC43, HKU1 and 229E) for 30 min. In the Monte Carlo simulations, the viral shedding was a stochastic variable following the distribution log_10_(*E*_*virus*_)∼*Normal*(4, 0.5). The half-life period of the virus also followed the triangular distribution with the lower limit, upper limit and mode of log_10_(0.5), log_10_(100) and log_10_(10) based on the data reported in the literature^8^. The deposition velocities were estimated following the method proposed by Lai et al. ^10^ based on the size distribution ^11^ of particles generated by breathing and coughing. For the exposure estimation, the total respiratory volume was assumed to be 0.6 m^3^ per hour and the deposition of particles in the respiratory system was estimated by lung deposition model for bioaerosols^12^.

The results indicated that the median infection risk to contract COVID-19 via the aerosol route was about 3.7×10^−5^ (95% confidence interval: 3.5×10^−6^ to 4.4×10^−4^) for one hour of exposure, which was more than three orders of magnitude lower than the risk due to close contact (about 12.8% within 1 m) based on the meta-analysis ^2^. It is reasonable to assume that 5 m is far enough to prevent the transmission routes of close contact (direct/indirect) or large respiratory droplets, so the infection risk is dominated by aerosol transmission. An additional intermediate data point from the meta-analysis ^2^ was that the infection risk was about 2.6% when the distance was > 1m. The above results suggested that the infection risk exponentially decreased with the increasing distance from the infected individual in the room. A simple exponential regression model *Risk*=10^−0.90×*D*+0.10^ (*D*<=5 m) could be utilized to roughly estimate the magnitude of infection risk for the considered scenario (Figure 1), where *D* is the distance from the infected individual.

**Figure 1.**
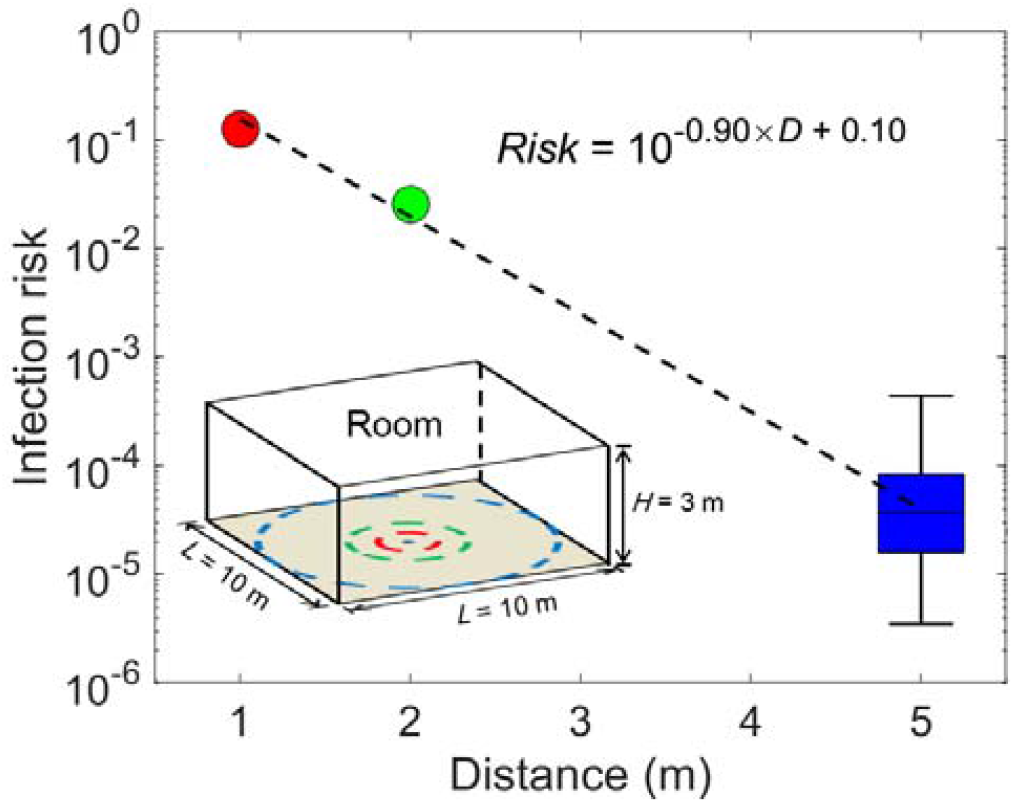
Infection risk at different distances from an infected individual in a room with the size of 10 m (width)×10 m (length)×3 m (height) for one hour exposure.

Larger rooms could reduce the infection risk via aerosol by enhancing the dilution of the airborne virus-laden particles. The decreasing rate of the risk was correlated with the area of the rooms (*L*^2^) as shown in Figure 2. The median risk decreased by about a factor of 4, from 3.7×10^−5^ to 1.0×10^−5^, when the room size increased from 10 m to 20 m. The infection risk decreased slower for further distance (*D*>5 m) than that within 5 m. The ventilation rates had moderate effects on the infection risk due to the modulation of deposition. The median risk increased from 3.7×10^−5^ with 1 ACH to 7.0×10^−5^ with 0.1 ACH as shown in Figure 3.

**Figure 2.**
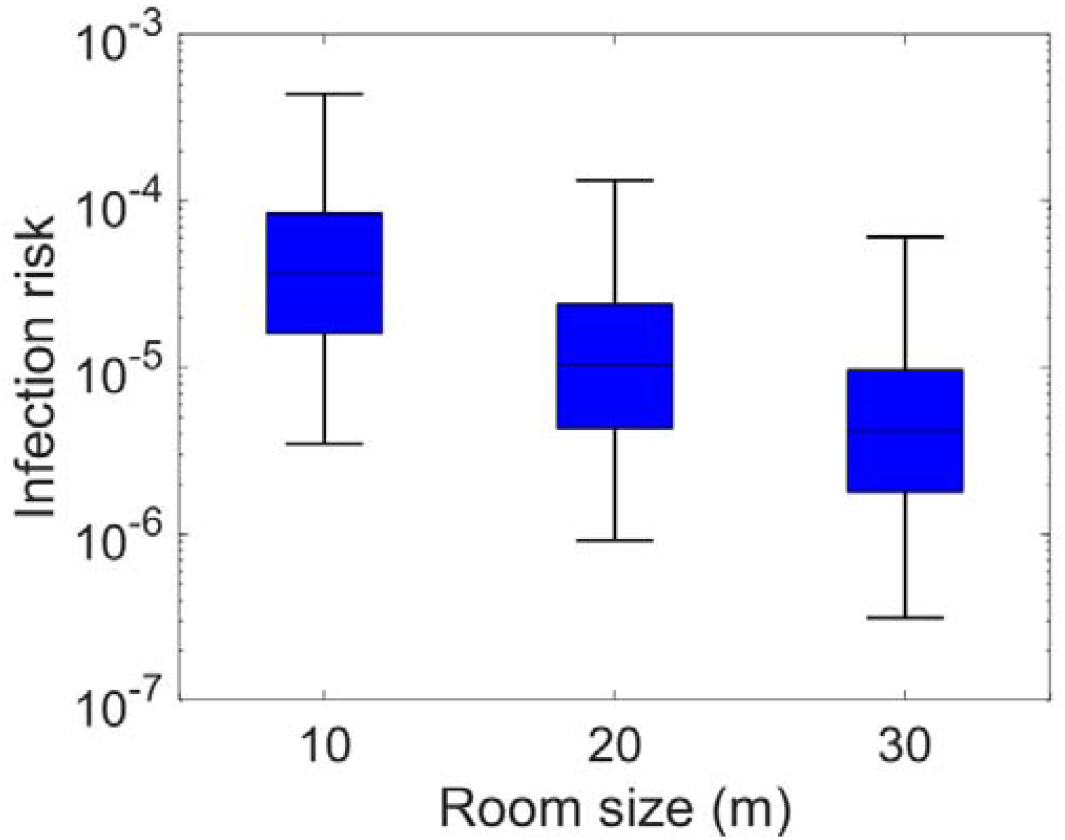
Infection risk via aerosol transmission in rooms of various sizes with one infected individual for one hour exposure.

**Figure 3.**
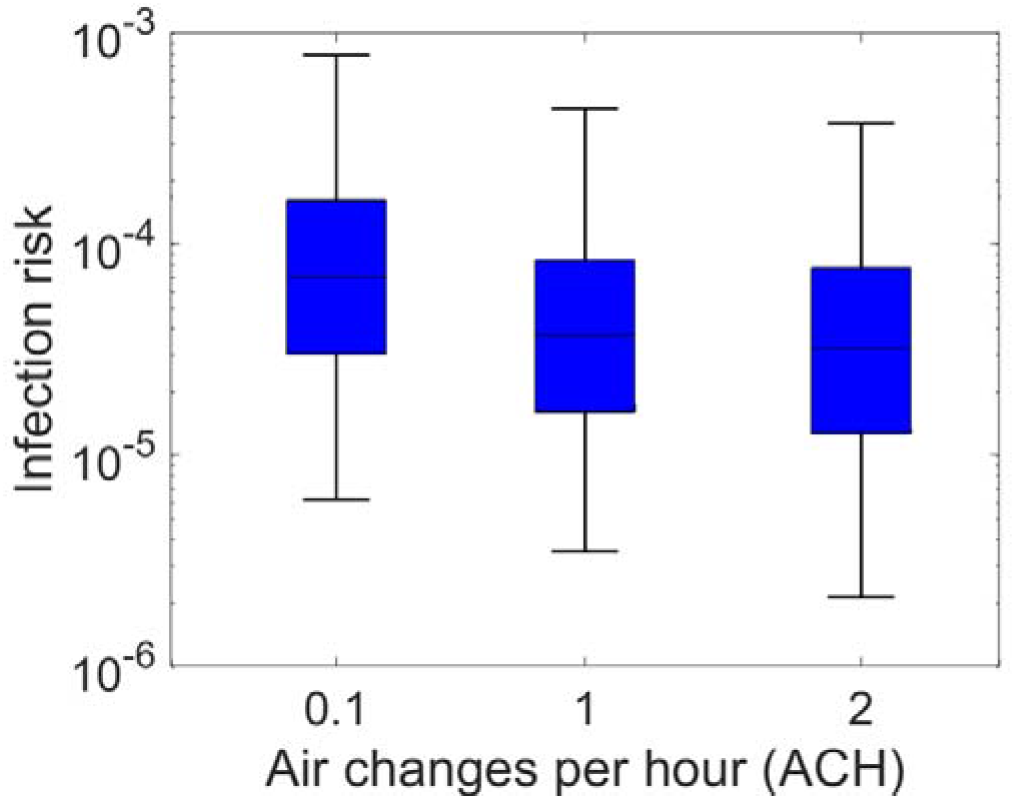
Infection risk via aerosol transmission in a room with the size of 10 m (width)×10 m (length)×3 m (height) with different ventilation rates for one hour exposure.

The recently developed dose-response relation could be utilized to quantify the magnitude of the infection risk via aerosol transmission. The uncertainties in the estimations were mainly due to the dose-response relation and the viral shedding. Despite the uncertainties, the assessment could still improve the understanding of the relative importance of different transmission routes. The infection risk via aerosol transmission with one-hour exposure (10^−6^ to 10^−4^) was more than three orders of magnitude lower than the risk caused by close contact (10^−2^ to 10^−1^).

Keeping distance is not enough to prevent or reduce the exposure to aerosol indoors. Wearing masks is helpful. The infection risk via aerosol transmission could increase for prolonged exposure duration, e.g. at home and office. In addition, the expected number of infected cases could be considerable if there are a large amount of exposed individuals, e.g. in clubs. As a result, it is necessary to be cautious for the potential aerosol transmission risk in such situations.

## Data Availability

All the data referred to in the manuscript are from published studies.

## Appendix

## Notes

### Competing Interest Statement

The authors have declared no competing interest.

### Funding Statement

No external funding was received for this study.

### Author Declarations

No ethical approval was needed for the study.

